# Clinical characterization and chest CT findings in laboratory-confirmed COVID-19: a systematic review and meta-analysis

**DOI:** 10.1101/2020.03.05.20031518

**Authors:** Golnaz Vaseghi, Marjan Mansourian, Raheleh Karimi, Kiyan Heshmat-Ghahdarijani, Sadegh Baradaran Mahdavi, Amirhossein Pezeshki, Behrooz Ataei, Alireza Zandifar, Omid Shafaat, Shaghayegh Haghjoo Javanmard

## Abstract

**Background:** Imagery techniques have been used as essential parts of diagnostic workup for patients suspected for 2019-nCoV infection, Multiple studies have reported the features of chest computed tomography (CT) scans among a number of 2019-nCoV patients.

**Method:** Study Identification was carried out in databases (PubMed, Embase and Cochrane Library) to identify published studies examining the diagnosis, the 2019 novel coronavirus (2019-nCoV). Heterogeneity among reported prevalence was assessed by computing p-values of Cochrane Q-test and I^2^ -statics. The pooled prevalence of treatment failure was carried out with a fixed effects meta-analysis model, generating the pooled 95% confidence interval. A random-effect model was used to pool the results since this model could incorporate the heterogeneity of the studies and therefore proved a more generalized result.

**Results:** According to the combined results of meta-analysis, the total 55% of corona patients were males. The mean age of the patients was 41.31 (34.14, 48.47). Two prevalent clinical symptoms between patients were fever, cough with prevalence of 85%, and 62%, respectively. Either Ground Glass Opacity GGO or consolidation was seen in 86% but 14% had **NO GGO or consolidation**.

The other rare CT symptoms were pericardial effusion, and pleural effusion with 4, 5, 7% prevalence, respectively. The most prevalent event was Either GGO or consolidation in 85% of patients.

**Conclusion:** The most CT-scan abnormality is Either Ground Glass Opacity GGO or consolidation however in few patients none of them might be observed, so trusting in just CT findings will lead to miss some patients.

## Introduction

The first cases of unusual pneumonia in early December of 2019 reported in Wuhan, China (1). Within 4 weeks, the pathogen identified as a novel betacorona virus named Severe Acute Respiratory Syndrome Coronavirus 2 (SARS-CoV2) or 2019-nCoV that mostly affects type II alveolar cells of human lungs (2). The virus uses a glycosylated spike protein and binds to Angiotensin-Converting Enzyme 2 (ACE2) Receptor that is similar to SARS-CoV virus (3). The incubation period is 1-14 days and it seems that asymptomatic patients can transmit the disease (4). The clinical manifestations include fever (83-98% of cases), dry cough (76-83% of cases), fatigue and myalgia (11-44% of cases). Also, other symptoms like headache, sore throat, abdominal pain and diarrhea has been reported in some cases. Laboratory tests showed lymphopenia (70%), increased Prothrombin Time (58%) and increased LDH (40%). The RT-PCR of the virus genome is still the standard diagnostic tool to confirm the disease (3).

Imagery techniques have been used as essential parts of diagnostic workup for patients suspected for 2019-nCoV infection (5). Multiple studies have reported the features of chest computed tomography (CT) scans among a number of 2019-nCoV patients (5-8). Findings such as ground glass opacities or mixed ground glass opacities and consolidations are of common reported features (9). However, the other related patterns and features of Chest CT such as the pulmonary lobes involved, remain to be better documented for 2019-nCoV infection.

Due to lack of approved treatment or vaccine, the attention has now been focused on the public health strategies to prevent the transmission of the new corona virus 2019 in the community (10). Therefore, in epidemic areas, it is crucial to early diagnose the patients highly suspected for 2019-nCoV according to epidemiological history and clinical presentation. The use of laboratory tests for 2019-nCoV may be time consuming or even limited by the lack of supply test kits. There is report of 5 patients with initially negative RT-PCR tests and positive chest CTs that eventually confirmed as 2019-nCoV infection via repeated swab tests (11). Furthermore, it has been proposed that in comparison with initial RT-PCR, chest CT had higher sensitivity to detect patients with 2019-nCoV. In this study the sensitivity of chest CT in suggesting 2019-nCoV was 97% based on positive RT-PCR results. The authors concluded that chest CT maybe considered as primary tool for detection of 2019-nCoV(12).

In this review, we systematically investigated the key features of chest CT findings of patients with confirmed diagnosis of 2019-nCoV infection. According to quick spread of 2019-nCoV (13) these CT characteristics may prompt the physicians suggest the diagnosis of 2019-nCoV as soon as indicated.

## Method

### Study Identification and Selection

A systematic search was carried out in databases (PubMed, Embase and Cochrane Library) to identify published studies examining the diagnosis, the 2019 novel coronavirus (2019-nCoV), in accordance with the Preferred Reporting Items for Systematic Reviews and Meta-Analyses (PRISMA) guidelines. There were two independent reviewers 2019-nCoV, respectively. We used “2019 Novel coronavirus”, “Wuhan virus”, “covid-19” and “CT-scan”, “Computed tomography” to identify the relevant studies. For the 2019-nCoV, we searched for all studies published in all language between till 20 February 2020. We used English and Chinese studies.

### Statistical analysis

Data on study design, sample size, study population and publication year were extracted in Microsoft Excel format, and then analysis was carried out using Stata software (version 13.1, Stata Corp, College Station, TX, USA). Heterogeneity among reported prevalence was assessed by computing p-values of Cochrane Q-test and I^2^ -statics (14). The pooled prevalence of treatment failure was carried out with a fixed effects meta-analysis model, generating the pooled 95% confidence interval. A random-effect model was used to pool the results since this model could incorporate the heterogeneity of the studies and therefore proved a more generalized result. With regard to the continuous variable presented in this meta-analysis, the standardized mean difference (SMD) and 95% confidence interval (CI) were computed using the random effects model. The SMD and 95% CI represent the pooled results of this study. Publication bias was assessed by Egger’s and Beggs’ tests at 5% significant level (15). Point prevalence, as well as 95% confidence intervals, was presented in the forest plot format for categorical variables (appendix). In this plot, the size of each box indicated the weight of the study, while each crossed line refers to 95% confidence interval. The reported values were two-tailed, and hypothesis testing results were considered statistically significant at *p* = 0.05.

## Results

### Literature search and study identification

The flowchart of database search was summarized in Fig. 1. Overall, 16 studies were identified after the initial database search, and 7 were excluded based on titles and abstracts mainly because they were irrelevant to the study purpose. The 9 remaining studies underwent full-text review.

**Figure 1.**
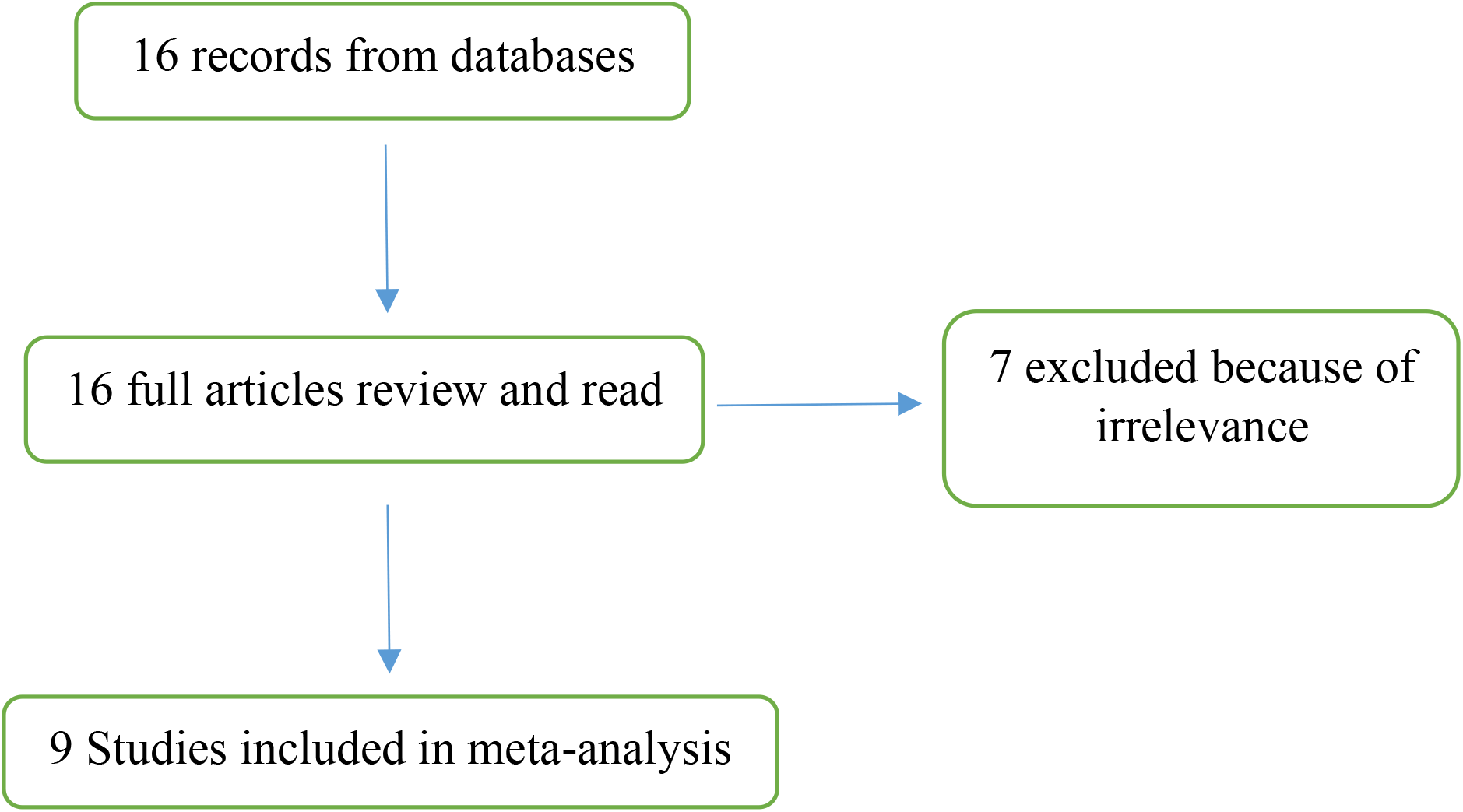
Flowchart representing the selection process

### Meta-analysis for the different clinical and CT-scan symptoms in Corona patients

Nine datasets with 293 patients from the different data bases were included in meta-analysis. Significant heterogeneity was detected (p for Cochrane’s Q test <0.001). Meta-analysis with random-effect model or fixed effect model if appropriates are showed in tabel.1.

The results based on clinical and CT-scan symptoms are presented in table.1. According to the combined results of meta-analysis, the total 55% of corona patients were males. The mean age of the patients was 41.31 (34.14, 48.47). Two prevalent clinical symptoms between patients were fever, cough with prevalence of 85%, and 62%, respectively. The rare CT-scan symptoms were lymphadenopathy (LAP), pericardial effusion, and pleural effusion with 4, 5, 7% prevalence, respectively. Bilateral involvement of lung was observed in 82% of patients (Figure.2).

**Table 1:**
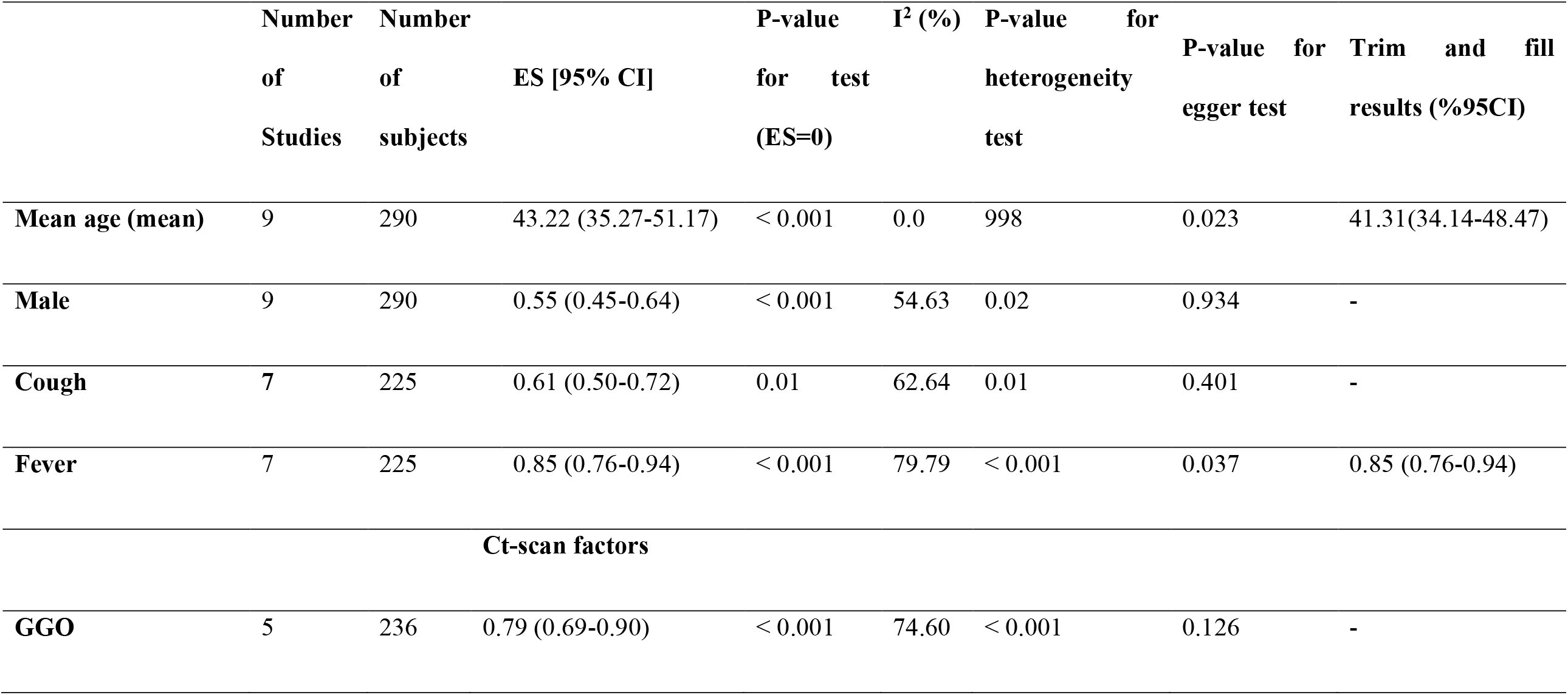
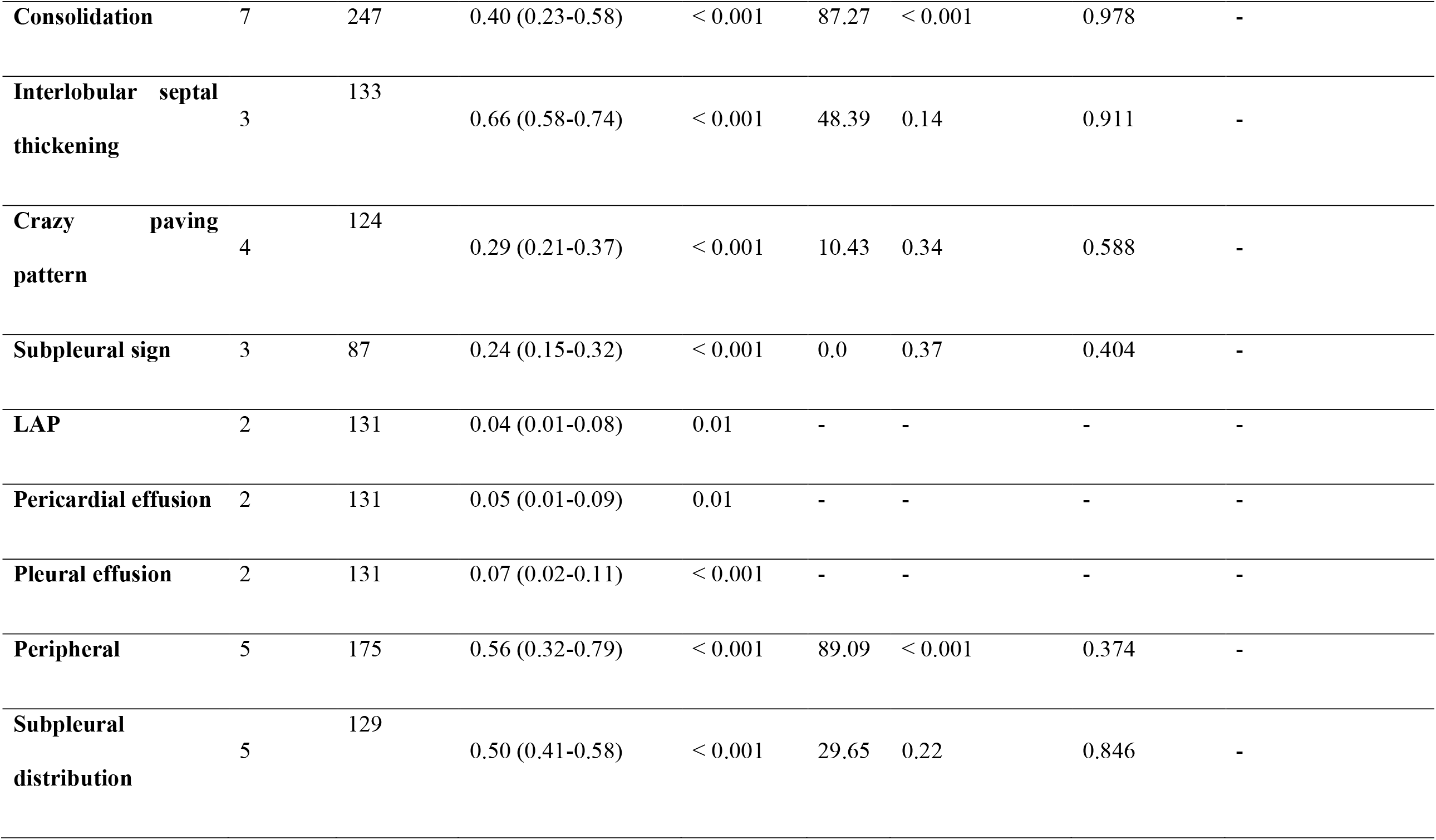
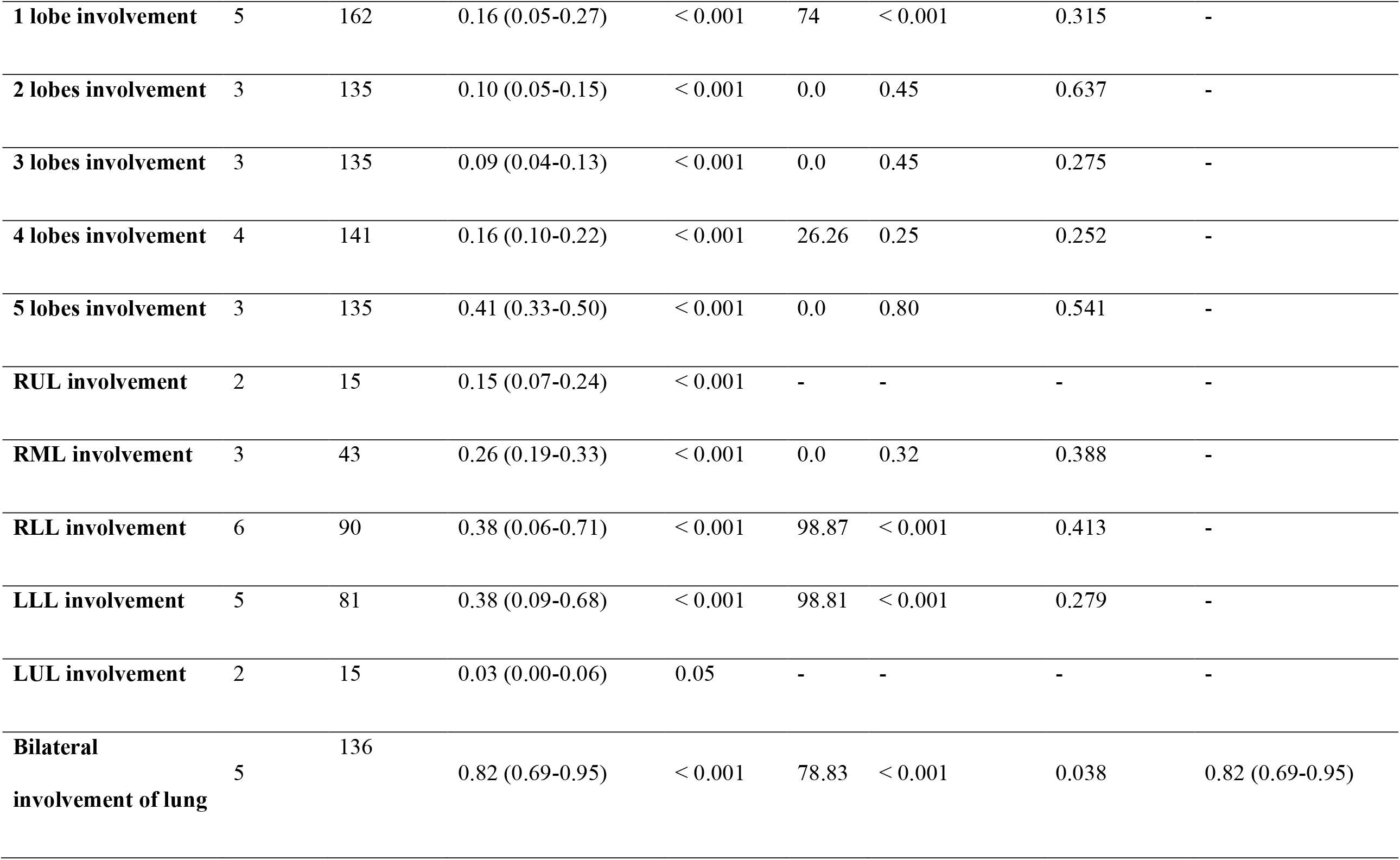
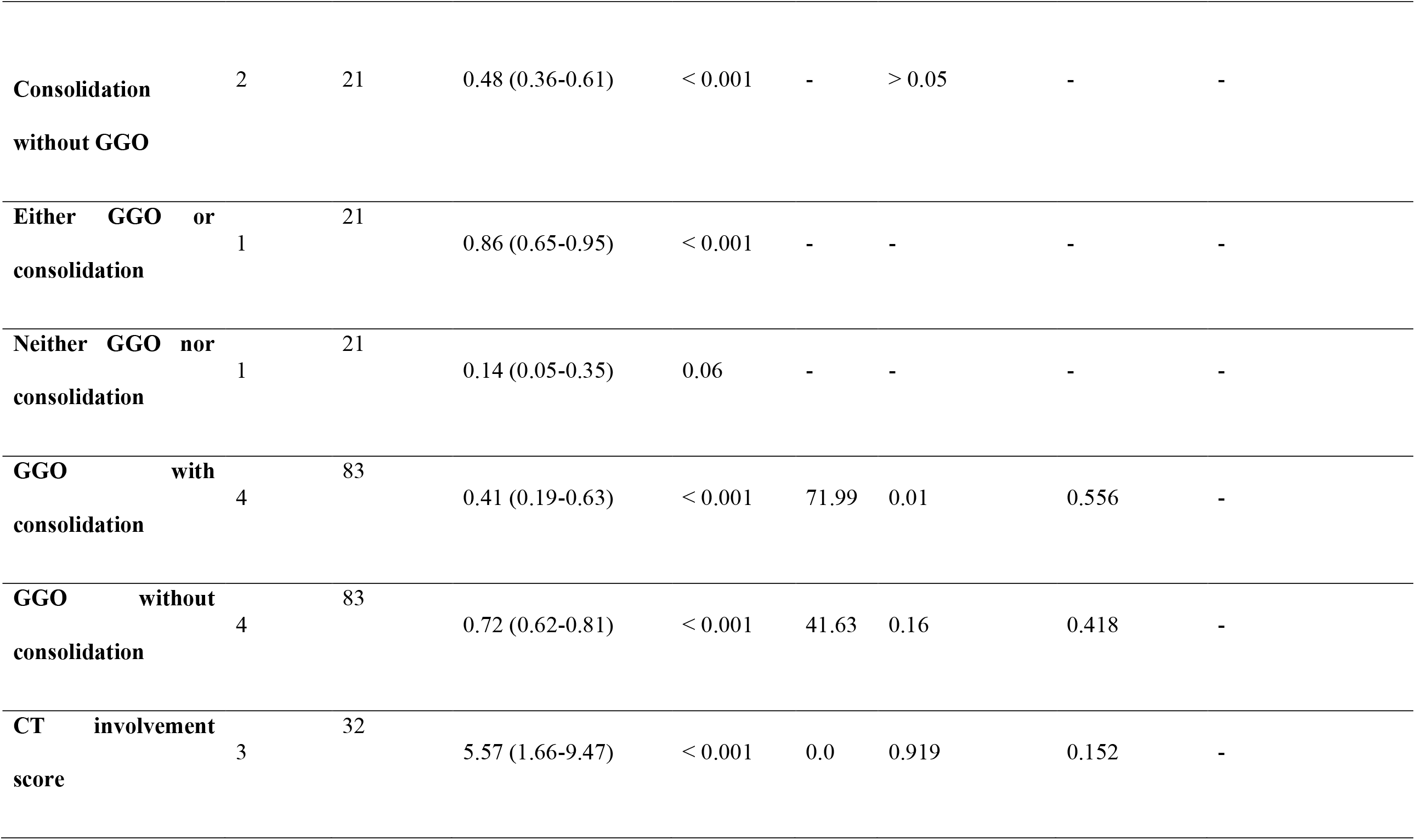
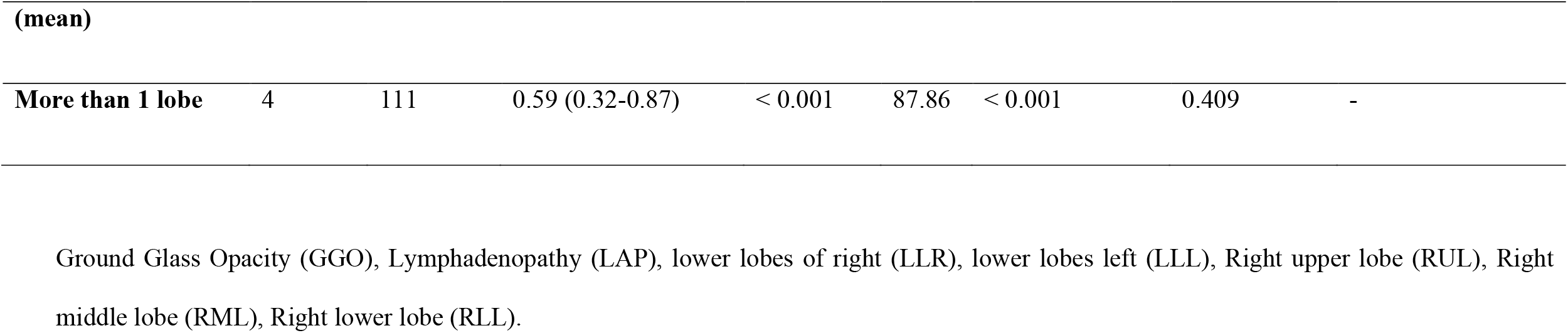
Summery of the study characteristics and results of meta-analysis statistics.

**Figure 2:**
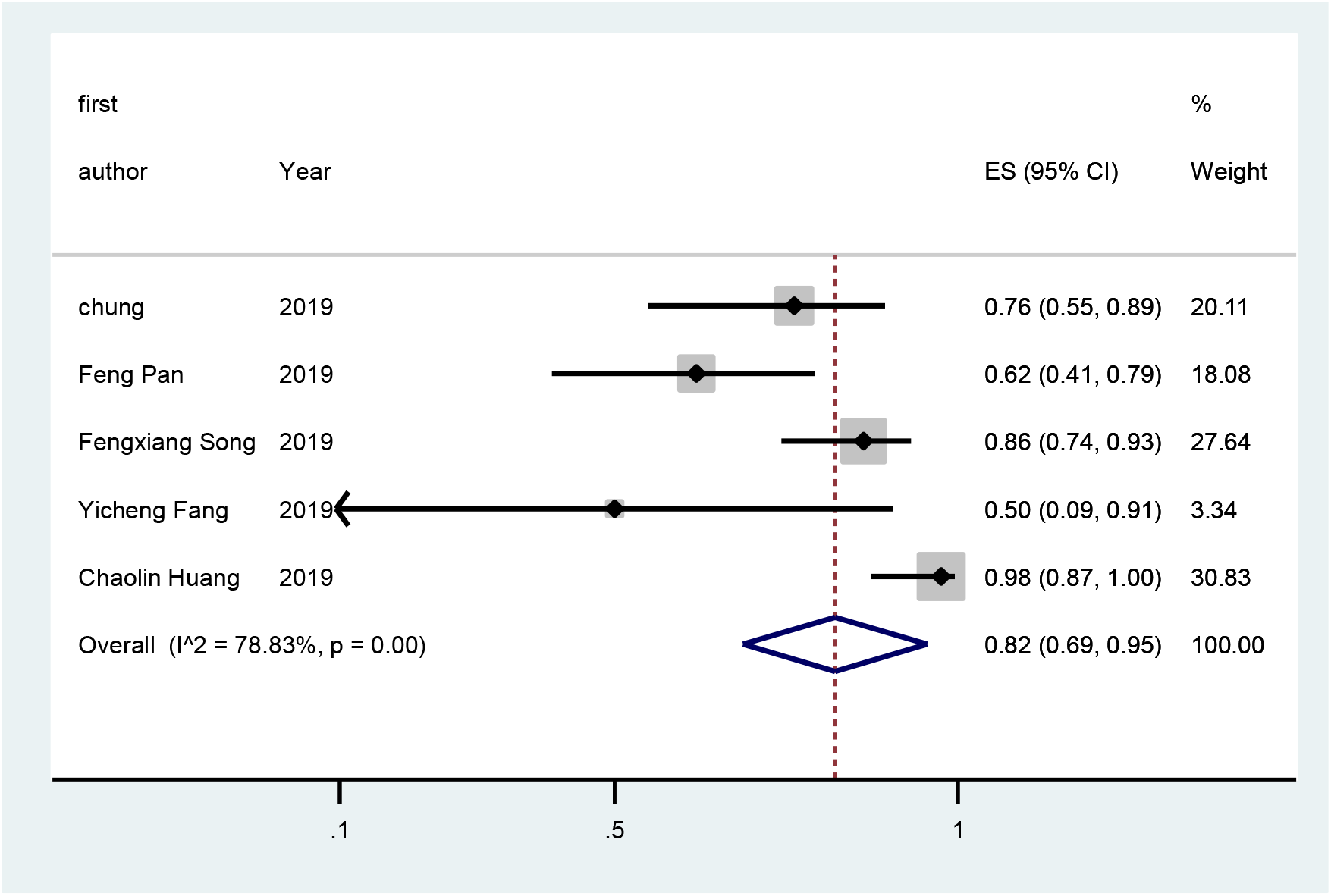
Forest plot of overall proportion of bilateral involvement of lung in Corona patients

Total, 7 studies presented consolidation in 40% of Corona subjects (Figure. 3), and 5 studies CT-scan results showed GGO status in about 80% of patients (Figure.4). Either GGO or consolidation (86%), and neither GGO, nor consolidation (14%) status reported only in a study with 21 subjects. The GGO with consolidation, and GGO without consolidation reported in 4 studies with 83 sample size in which GGO without consolidation was more prevalent (72%) than the other one.

**Figure 3:**
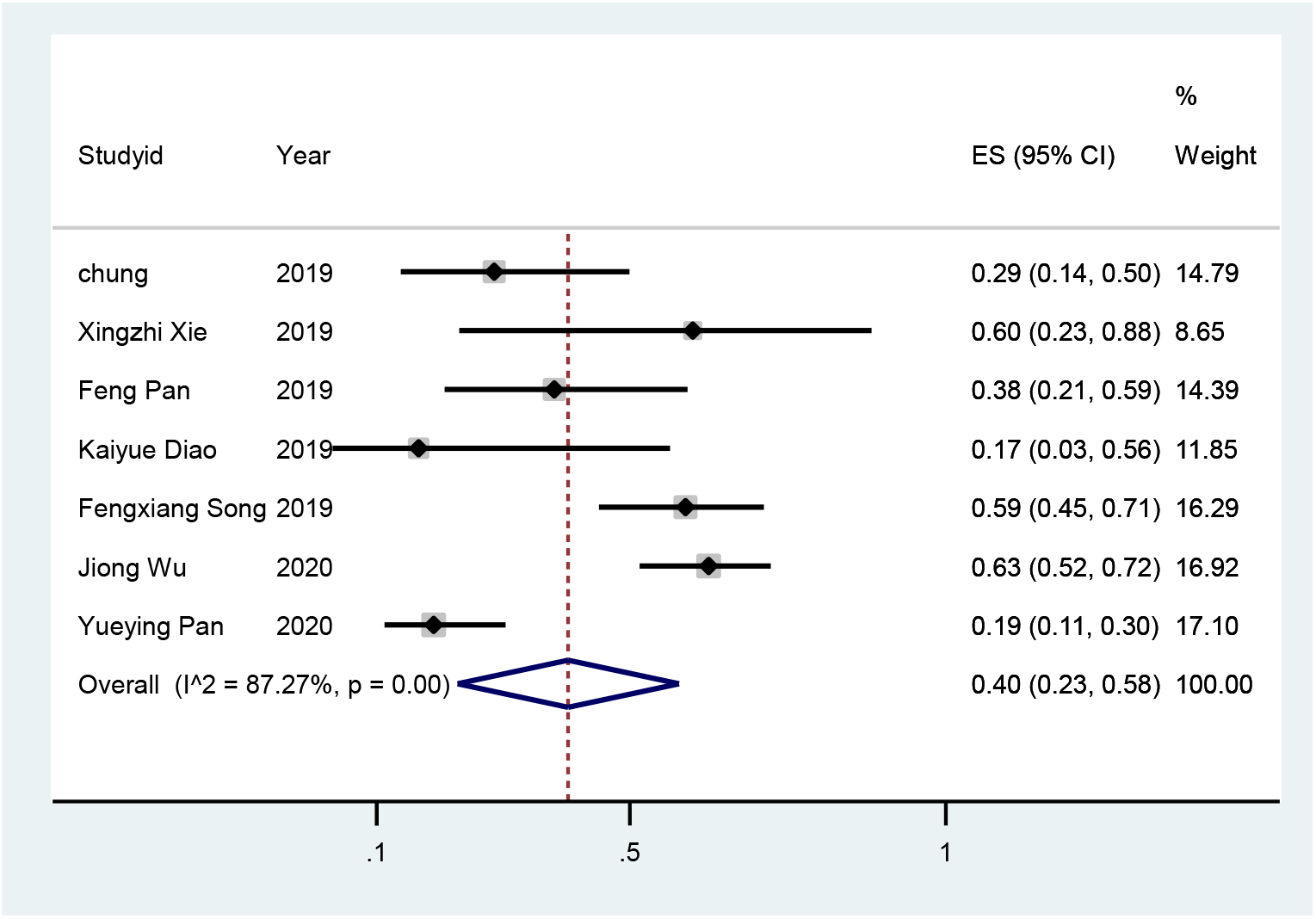
Forest plot of overall proportion of consolidation status in Corona patients

**Figure 4:**
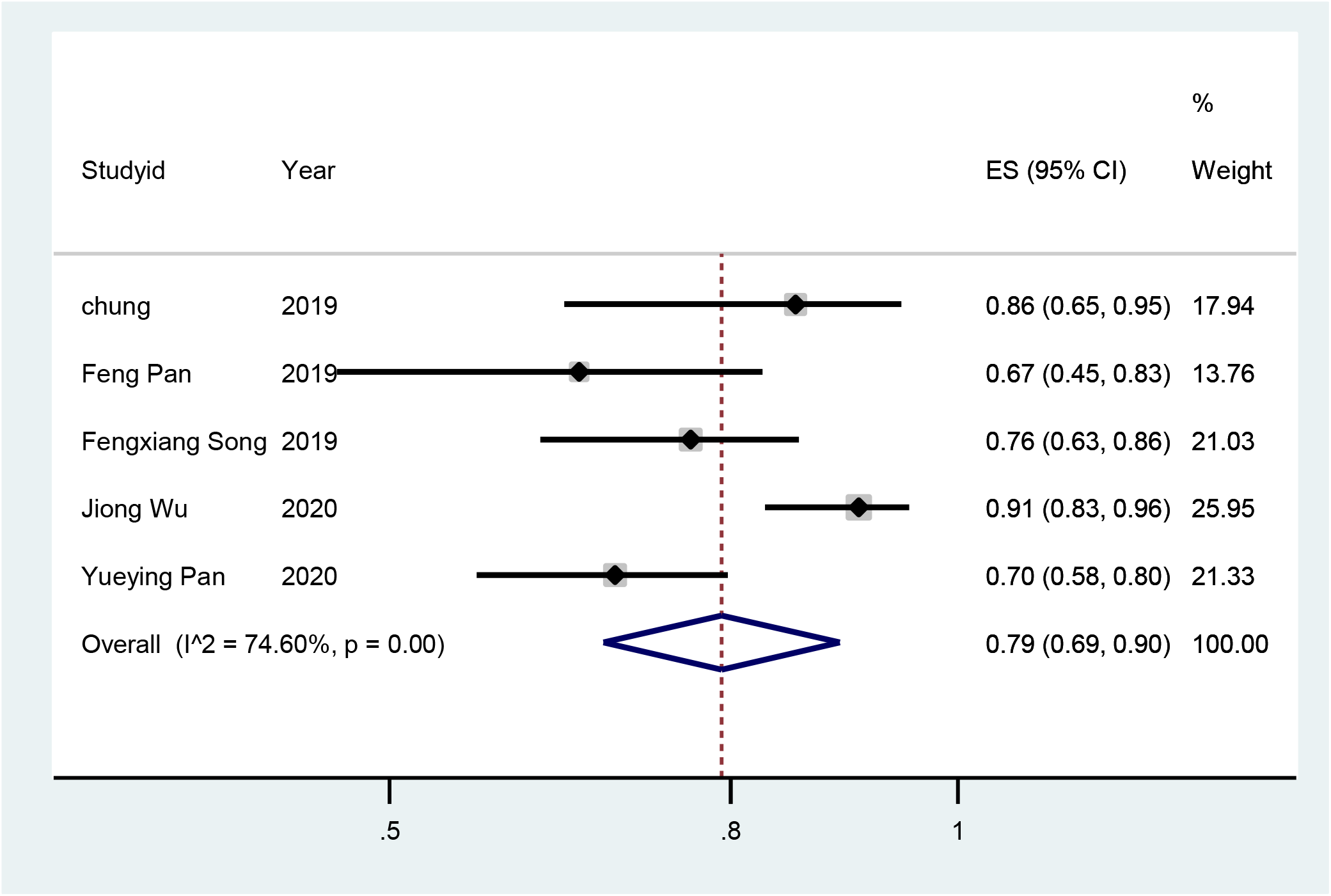
Forest plot of overall proportion of GGO status in Corona patients

### Publication bias

The funnel plots for the meta-analysis of the prevalence of different symptoms in Corona patients were not shown. The plots were symmetrical on visual inspection, indicating low risk of publication bias in most of status. The Egger’s regression tests also demonstrated in Table.1. For studies with high risk of publication the adjusted trim and fill statistics were presents in table.1

### Grading the quality of evidence

The quality of evidence and confidence for the main outcomes (primary outcomes) of including studies in our review will be evaluated and assessed on the basis of the Grading of Recommendations Assessment, Development and Evaluation (GRADE) guideline (16). The quality of evidence will be adjudicated into 4 levels: “very low”, “low”, “moderate” or “high” judgment. Any discrepancy will be resolved by consensus or consultation with a third review author (RK). All studies had the high score in judgment.

According to rapid spread of new corona virus 2019 in the world (reported in 86 countries as of March 5, 2020), it is crucial to stablish robust clinical data to help health care providers control the outbreak (17). Early diagnosis of 2019-nCoV can lead to patient isolation strategies which in turn leads to reduction of viral transition. The epidemiological history may be vague and the sign and symptoms of the disease may resemble other viral infections. The RT-PCR tests of serum or respiratory specimens may be unavailable for all suspected patients as well. There is the possibility of initial false negative results of RT-PCR tests due to lack of replicable nucleic acid or technical errors as well (18). In this context, chest CT images are important elements of clinical workflow of 2019-nCoV, with respect to screening, diagnosis and follow-up. In this systematic review and meta-analysis, we aimed to find the answer for this question that whether there is any fix pattern of pulmonary involvement in chest CT scans of patients with 2019-nCoV infection according to available evidence.

Our pooled data revealed that the most common patterns of CT scan in infected patients were Ground Glass Opacity (GGO), interlobular septal thickening and consolidation (presented in 79%, 66% and 40% of cases respectively) especially in bilateral lower lobes of lung (reported in 38% of patients). Right middle lobe, right upper lobe and left upper lobe are involved less than lower lobes (26%, 15% and 3% respectively). The patterns of lesions are mostly bilateral (82%) and peripheral (56%). Crazy paving pattern, LAP, pericardial and pleural effusion and sub-pleural signs were the other CT scan findings accordingly (6, 19, 20). However there are positive RT-PCR cases with normal pattern of chest CT scan suggesting not to exclude these patients of 2019-nCoV (21).

Above findings are in consistent with recent studies in which the chest CT characteristics of 2019-nCoV pneumonia were identified. Bilateral peripheral ground-glass and consolidative pulmonary opacities were found in CT scans of 21 patients from China. In this study, 15 cases (71%) had the involvement of more than 2 lobes. Also pulmonary nodules, cavitation, pleural effusions, and lymphadenopathy were absent in the cases (5). Another study among 6 patients with confirmed diagnosis of 2019-nCoV in China revealed multifocal or unifocal involvement of ground-glass opacity (GGO) with consolidation and fibrosis as basic pattern. In this study pleural effusion and lymphadenopathy were absent as well. Also the lesions generally progressed in follow-up CT scans obtained between 2 to 6 days after initial images (22).

It is worthwhile to consider the dynamic nature of chest CT findings of 2019-nCoV, i.e., the alteration of finings that may be observed in repeated measures. In a study among 21 non-respiratory compromised patients with 2019-nCoV, multiple CT images were performed in the process of recovery. The authors classified 4 radiologic stages. In stage 1 (0-4 days after onset of disease), GGO in the lower lobes was the main finding. In stage 2 (5-8 days after the onset) the infection extended and diffuse GGO, crazy-paving pattern and consolidation were observed. In stage 3 (9-13 days after the onset, peak stage), the consolidation was the dominant abnormality and finally in sage 4 (≥14 days after the onset, absorption stage), consolidation was absorbed. There was no crazy-paving pattern but still a remarkable GGO abnormality in this stage. Furthermore, in this study, GGO and peripheral distribution were dominant (more than 50% of patients) in all 4 stages. Also, bilateral multilobe involvement was dominant in stages 2, 3 and 4. Similar to some case reports (23, 24), it can be concluded that chest CT findings of individuals with 2019-nCoV infection may be diverse and rapidly change during the disease course. Future research may focus on the main characteristics of the 2019-nCoV pulmonary lesions in different clinical stages and the correlation of these findings with the prognosis or outcome in the affected individuals.

In our study we observed neither consolidation nor GGO in 14% of patients, So, trusting only on CT scan findings leads to miss a few patients with confirmed laboratory finding of 2019-nCoV.

2019-nCoV, SARS-CoV and MERS are subtypes of a common family. The comparison of radiologic features of pulmonary involvement of these viral pathogens with other viruses, may help radiologists make more accurate diagnoses based on CT images. Generally, the chest CT scan findings of the 2019-nCoV, SARS-CoV and MERS are similar (25, 26), however 2019-nCoV may present with specific characteristics. CT scan in patients with influenza lower respiratory tract infection (LRTI) shows diffused GGO and localized bronchial wall thickening in upper and mid lobes in comparison with 2019-nCoV that mostly affects lower lobes (21, 27, 28). GGO and bronchial wall thickening in mid and upper lobes and centrilobular opacities are radiologic manifestations of RSV LRTI and bronchiolitis but the consolidation and GGO can be present in severe cases(29-31). SARS-CoV infection is another LRTI that appears with progressive pneumonia shows GGO in early phase of the disease and gradually turns into reticular opacities within 4 weeks. The pattern remains in CT scan for months especially in patients with prolonged dyspnea. The CT scan patterns of viral LRTI are not characteristic but the epidemic parameters in addition to signs and symptoms can lead us to the diagnosis (32-34).

In summary, bilateral peripheral pattern of GGO, interlobular septal thickening and consolidation mostly in bilateral lower lobes of lung in chest CT scans of suspected individuals in combination with epidemiological data, presence of fever and cough and laboratory positive results, are highly in favor of the diagnosis of 2019-nCoV in an individual, but this pattern does not observed in all cases as we showed 14% with laboratory finding may not have either consolidation or GGO which may be missed during screening with CT scan.

## Data Availability

I accept the free availability of our manuscript.

## Notes

### Competing Interest Statement

The authors have declared no competing interest.

### Funding Statement

This study was supported by the Isfahan University of Medical Sciences.

## References

1. Huang C, Wang Y, Li X, Ren L, Zhao J, Hu Y, et al. Clinical features of patients infected with 2019 novel coronavirus in Wuhan, China. The Lancet. 2020;395(10223):497–506.

2. Zhu N, Zhang D, Wang W, Li X, Yang B, Song J, et al. A novel coronavirus from patients with pneumonia in China, 2019. New England Journal of Medicine. 2020.

3. Del Rio C, Malani PN. COVID-19-New Insights on a Rapidly Changing Epidemic. Jama. 2020.

4. Bai Y, Yao L, Wei T, Tian F, Jin D-Y, Chen L, et al. Presumed Asymptomatic Carrier Transmission of COVID-19. Jama. 2020.

5. Chung M, Bernheim A, Mei X, Zhang N, Huang M, Zeng X, et al. CT imaging features of 2019 novel coronavirus (2019-nCoV). Radiology. 2020:200230.

6. Wu J, Wu X, Zeng W, Guo D, Fang Z, Chen L, et al. Chest CT Findings in Patients with Corona Virus Disease 2019 and its Relationship with Clinical Features. Investigative radiology. 2020.

7. Fang Y, Zhang H, Xu Y, Xie J, Pang P, Ji W. CT manifestations of two cases of 2019 novel coronavirus (2019-nCoV) pneumonia. Radiology. 2020:200280.

8. Song F, Shi N, Shan F, Zhang Z, Shen J, Lu H, et al. Emerging coronavirus 2019-nCoV pneumonia. Radiology. 2020:200274.

9. Kanne JP. Chest CT findings in 2019 novel coronavirus (2019-nCoV) infections from Wuhan, China: key points for the radiologist. Radiology. 2020:200241.

10. Wu Z, McGoogan JM. Characteristics of and Important Lessons From the Coronavirus Disease 2019 (COVID-19) Outbreak in China: Summary of a Report of 72 314 Cases From the Chinese Center for Disease Control and Prevention. Jama.

11. Xie X, Zhong Z, Zhao W, Zheng C, Wang F, Liu J. Chest CT for typical 2019-nCoV pneumonia: relationship to negative RT-PCR testing. Radiology. 2020:200343.

12. Ai T, Yang Z, Hou H, Zhan C, Chen C, Lv W, et al. Correlation of Chest CT and RT-PCR Testing in Coronavirus Disease 2019 (COVID-19) in China: A Report of 1014 Cases. Radiology. 2020.

13. The epidemiological characteristics of an outbreak of 2019 novel coronavirus diseases (COVID-19) in China]. Zhonghua liu xing bing xue za zhi = Zhonghua liuxingbingxue zazhi. 2020;41(2):145–51.

14. Rucker G, Schwarzer G, Carpenter JR, Schumacher M. Undue reliance on I(2) in assessing heterogeneity may mislead. BMC medical research methodology. 2008;8:79.

15. Borenstein M, Hedges LV, Higgins JP, Rothstein HR. A basic introduction to fixed-effect and random-effects models for meta-analysis. Research synthesis methods. 2010;1(2):97–111.

16. Guyatt GH, Oxman AD, Vist GE, Kunz R, Falck-Ytter Y, Alonso-Coello P, et al. GRADE: an emerging consensus on rating quality of evidence and strength of recommendations. BMJ (Clinical research ed). 2008;336(7650):924–6.

17. Xu B, Kraemer MUG. Open access epidemiological data from the COVID-19 outbreak. The Lancet Infectious diseases. 2020.

18. Fang Y, Zhang H, Xie J, Lin M, Ying L, Pang P, et al. Sensitivity of Chest CT for COVID-19: Comparison to RT-PCR. Radiology. 2020:200432.

19. Chen L, Liu HG, Liu W, Liu J, Liu K, Shang J, et al. [Analysis of clinical features of 29 patients with 2019 novel coronavirus pneumonia]. Zhonghua jie he he hu xi za zhi = Zhonghua jiehe he huxi zazhi = Chinese journal of tuberculosis and respiratory diseases. 2020;43(0):E005.

20. Chan JF-W, Yuan S, Kok K-H, To KK-W, Chu H, Yang J, et al. A familial cluster of pneumonia associated with the 2019 novel coronavirus indicating person-to-person transmission: a study of a family cluster. The Lancet. 2020;395(10223):514–23.

21. Yang W, Cao Q, Qin L, Wang X, Cheng Z, Pan A, et al. Clinical characteristics and imaging manifestations of the 2019 novel coronavirus disease (COVID-19): A multi-center study in Wenzhou city, Zhejiang, China. Journal of Infection. 2020.

22. Diao K, Han P, Pang T, Li Y, Yang Z. HRCT imaging features in representative imported cases of 2019 novel coronavirus pneumonia. Precision Clinical Medicine. 2020.

23. Duan Y-n, Qin J. Pre-and posttreatment chest CT findings: 2019 novel coronavirus (2019-nCoV) pneumonia. Radiology. 2020:200323.

24. Shi H, Han X, Zheng C. Evolution of CT manifestations in a patient recovered from 2019 novel coronavirus (2019-nCoV) pneumonia in Wuhan, China. Radiology. 2020:200269.

25. Ooi GC, Khong PL, Müller NL, Yiu WC, Zhou LJ, Ho JC, et al. Severe acute respiratory syndrome: temporal lung changes at thin-section CT in 30 patients. Radiology. 2004;230(3):836–44.

26. Das KM, Lee EY, Jawder SEA, Enani MA, Singh R, Skakni L, et al. Acute Middle East respiratory syndrome coronavirus: temporal lung changes observed on the chest radiographs of 55 patients. American Journal of Roentgenology. 2015;205(3):W267–S74.

27. Song F-x, Jun Z, Shi Y-x, Zhang Z-y, Feng F, Zhou J-j, et al. Bedside chest radiography of novel influenza A (H7N9) virus infections and follow-up findings after short-time treatment. Chinese medical journal. 2013;126(23):4440–3.

28. Kim M-C, Kim MY, Lee HJ, Lee S-O, Choi S-H, Kim YS, et al. CT findings in viral lower respiratory tract infections caused by parainfluenza virus, influenza virus and respiratory syncytial virus. Medicine. 2016;95(26).

29. Mayer J, Lehners N, Egerer G, Kauczor H, Heussel C, editors. CT-morphological characterization of respiratory syncytial virus (RSV) pneumonia in immune-compromised adults. RöFo-Fortschritte auf dem Gebiet der Röntgenstrahlen und der bildgebenden Verfahren; 2014: © Georg Thieme Verlag KG.

30. Syha R, Beck R, Hetzel J, Ketelsen D, Grosse U, Springer F, et al. Humane metapneumovirus (HMPV) associated pulmonary infections in immunocompromised adults— Initial CT findings, disease course and comparison to respiratory-syncytial-virus (RSV) induced pulmonary infections. European journal of radiology. 2012;81(12):4173–8.

31. Ko JP, Shepard J-AO, Sproule MW, Trotman-Dickenson B, Drucker EA, Ginns LC, et al. CT manifestations of respiratory syncytial virus infection in lung transplant recipients. Journal of computer assisted tomography. 2000;24(2):235–41.

32. Antonio GE, Wong KT, Tsui EL, Chan DP, Hui DS, Ng AW, et al. Chest radiograph scores as potential prognostic indicators in severe acute respiratory syndrome (SARS). American Journal of Roentgenology. 2005;184(3):734–41.

33. Müller NL, Ooi GC, Khong PL, Zhou LJ, Tsang KW, Nicolaou S. High-resolution CT findings of severe acute respiratory syndrome at presentation and after admission. American Journal of Roentgenology. 2004;182(1):39–44.

34. Paul NS, Roberts H, Butany J, Chung T, Gold W, Mehta S, et al. Radiologic pattern of disease in patients with severe acute respiratory syndrome: the Toronto experience. Radiographics. 2004;24(2):553–63.

